# Barriers and facilitators to diabetes management among adults and healthcare providers in a peri-urban Ugandan health facility: A qualitative study

**DOI:** 10.64898/2026.06.22.26356287

**Authors:** Larissa Ngazet Yangbalet Kezza, Ronald Kooko, David Musoke, Rogers Kisame, Aristide Desire Komangoya-Nzonzo, Olivia Nakisita, Marius Wilfried Wanikomane Dandy

**Author notes:** **Correspondence:** Larissa Ngazet Yangbalet Kezza. **Email addresses of authors:** Ronald Kooko, David Musoke, Rogers Kisame, Aristide Desire Komangoya-Nzonzo, Olivia Nakisita, Marius Wilfried Wanikomane Dandy.

## Abstract

Diabetes mellitus is an increasing public health challenge in Uganda and other low- and middle-income countries, where health systems face growing demands for chronic disease care. Although quantitative studies have documented poor glycemic control and health system constraints, less is known about how patients and healthcare providers experience diabetes management in peri-urban public health settings. This study explored barriers and facilitators to diabetes management among adults with diabetes mellitus and healthcare providers at a peri-urban health facility in Uganda. We conducted a qualitative descriptive study at Kasangati Health Centre IV, Wakiso District, Uganda, between February and March 2025. Data were collected through 15 in-depth interviews with adults living with diabetes mellitus and 8 key informant interviews with healthcare providers involved in diabetes care. Participants were purposively selected based on their experience with diabetes management and service delivery. Interviews were audio-recorded, transcribed verbatim, translated where necessary, and analyzed using a hybrid inductive–deductive thematic approach informed by the Theoretical Domains Framework. Five interrelated themes were identified: (1) institutional and environmental factors influencing access to diabetes care; (2) cognitive and informational factors influencing medication adherence; (3) social influences on diabetes management; (4) emotional experiences of patients and healthcare providers; and (5) self-management strategies and continuity of care. Across these themes, participants identified barriers including resource limitations, communication challenges, medication management difficulties, stigma, emotional distress, and weak follow-up systems. Facilitators included peer support, religious and community networks, health education, provider flexibility, and patient-developed adherence strategies. Diabetes management was influenced by interacting health-system, social, informational, and behavioural factors. Resource constraints, limited health literacy, stigma, and weak follow-up systems hindered effective management, while social support, health education, and patient self-management strategies facilitated continued engagement in care. Interventions that strengthen chronic care services, patient education, and community support may improve diabetes outcomes in similar resource-constrained settings.

## Introduction

The global burden of non-communicable diseases (NCDs) continues to rise, particularly in low-and middle-income countries (LMICs), where health systems have historically been oriented toward acute infectious diseases rather than chronic care. Diabetes mellitus (DM) is among the most significant NCDs globally, with an estimated 589 million adults aged 20–79 years living with the condition in 2025, nearly four-fifths of whom reside in LMICs [1]. Diabetes is associated with substantial morbidity, mortality, reduced quality of life, and increasing healthcare costs for individuals and health systems [1, 2].

Sub-Saharan Africa is experiencing a rapid increase in diabetes prevalence driven by urbanization, population ageing, physical inactivity, and dietary transitions [1, 3]. However, health systems in the region remain constrained by shortages of trained health workers, limited diagnostic capacity, weak supply chains, and inadequate long-term follow-up systems, contributing to delayed diagnosis, poor glycaemic control, and preventable complications [3, 4].

In Uganda, diabetes is becoming an important public health challenge. Although the 2014 WHO STEPwise Survey reported a prevalence of 1.4% among adults, more recent evidence suggests there is an increasing burden of diabetes and the associated risk factors, particularly in urban and peri-urban settings undergoing rapid lifestyle transitions [5, 6, and 7]. Previous studies in Uganda have documented substantial challenges in diabetes management, including poor glycaemic control, inadequate diabetes knowledge, medication non-adherence, financial barriers to care, health-system constraints such as shortages of medicines, diagnostic supplies, and trained healthcare workers, as well as the influence of social support, health literacy, and patient provider communication on diabetes self-management and treatment outcomes [8, 9, and 10]. However, much of this evidence has been generated through quantitative studies that describe the prevalence and correlates of diabetes-related outcomes, providing limited insight into how patients and healthcare providers experience and navigate diabetes management within routine care settings. The Theoretical Domains Framework (TDF) is a behavioural science framework that synthesizes determinants of behaviour into key domains and is widely used in implementation research to identify barriers and facilitators to health-related practices [11]. By providing a structured approach to understanding behavioural influences, the framework can support the identification of contextually appropriate interventions for strengthening diabetes care. Therefore, this study explored barriers and facilitators to diabetes management among adult patients and healthcare providers attending Kasangati Health Centre IV in Wakiso District, Uganda. Guided by the TDF, the study sought to identify behavioural and health-system factors influencing diabetes management to inform contextually appropriate interventions for strengthening diabetes care in similar settings.

## Materials and Methods

### Ethical considerations

The study was conducted in accordance with the Declaration of Helsinki and approved by the Makerere University School of Public Health Research and Ethics Committee (MakSPH-REC_502). Administrative permission was obtained from Kasangati Health Centre IV (HCIV).

Informed verbal consent was obtained from all participants before data collection. The use of verbal consent was approved by the Research Ethics Committee to minimize the collection of identifying information and enhance confidentiality. Participants received information about the study objectives, procedures, potential risks and benefits, voluntary participation, and confidentiality protections in their preferred language (*Luganda* for patients and English for healthcare providers). They were informed of their right to decline participation, refuse to answer any question, or withdraw from the study at any time without consequences.

No personal identifiers were collected. Audio recordings and transcripts were assigned unique identification codes, and all identifying information was removed during transcription. Electronic data were encrypted and stored on password-protected devices accessible only to the research team.

### Study design and setting

This qualitative descriptive study was informed by the Theoretical Domains Framework (TDF) and explored barriers and facilitators to diabetes management from the perspectives of patients and healthcare providers. A qualitative descriptive approach was chosen to provide an in-depth understanding of participants’ experiences while generating findings relevant to healthcare practice and policy.

The study was conducted at Kasangati HCIV, a public health facility located in Wakiso District, approximately 16 km north of Kampala, Uganda. The facility serves a rapidly growing peri-urban population and provides a range of services, including outpatient care, maternal and child health services, emergency care, and management of non-communicable diseases. Kasangati HCIV was selected because of its large chronic care clinic and increasing demand for diabetes services within a primary healthcare setting.

### Participant selection and recruitment

A purposive sampling strategy was used to recruit participants with experience in diabetes management and service delivery. Twenty-three participants were enrolled, comprising 15 adults living with diabetes mellitus who participated in in-depth interviews (IDIs) and 8 healthcare providers who participated in key informant interviews (KIIs).

Patients were recruited from the chronic care clinic during routine clinic visits. Eligible participants were aged 18 years or older, had been diagnosed with diabetes mellitus and receiving care at the facility for at least six months. Individuals with severe cognitive impairment, acute illness, or conditions that could limit meaningful participation were excluded.

Healthcare providers were selected to capture diverse perspectives across the diabetes care pathway and included clinical officers, nurses, a laboratory technician, a pharmacist/dispenser, and a facility administrator. Providers were required to have worked at the facility for at least six months and to be directly involved in diabetes-related service delivery or management.

### Development of interview guides and data collection

Separate semi-structured interview guides were developed for patients and healthcare providers. The guides were informed by relevant TDF domains, including Knowledge, Skills, Social Influences, Environmental Context and Resources, Behavioural Regulation, Beliefs about Consequences, and Emotion. The guides were reviewed by the research team and piloted at Goma Health Centre IV, a facility with characteristics similar to the study setting. Feedback from the pilot informed revisions to improve clarity, sequencing, and contextual relevance.

Data were collected through face-to-face interviews conducted between 21 February and 21 March 2025. Patient interviews were conducted in *Luganda*, while healthcare provider interviews were conducted in English. Interviews were held in private rooms within the facility, lasted approximately 45–60 minutes, and were audio-recorded with participant permission. Field notes were taken during and immediately after interviews to document contextual observations and support data interpretation.

### Saturation, research team, and reflexivity

Data collection and analysis occurred concurrently to facilitate assessment of thematic saturation. Saturation was evaluated through ongoing review of interview transcripts and regular debriefing meetings between the Principal Investigator (PI) and the research assistant. Initial code saturation among patients was observed after the twelfth interview, and three additional interviews were conducted to confirm thematic completeness where no new code emerged. Provider interviews continued until no substantially new perspectives emerged across the selected professional cadres. Data collection was conducted by a bilingual female research assistant with a Bachelor of Science in Public Health and three years of experience in qualitative research. She was fluent in English and *Luganda*, received training on the study protocol, research ethics, and qualitative interviewing techniques, and had no prior relationship with participants. Participants were informed of her independence from Kasangati HCIV to encourage open discussion and reduce social desirability bias. The PI has a background in public health and health systems research. To minimize the influence of pre-existing assumptions, analysis began with inductive coding before findings were interpreted through the TDF. Reflexive discussions were conducted throughout the study to examine emerging interpretations and potential sources of bias.

### Data analysis

Audio-recorded interviews were transcribed verbatim. *Luganda* interviews were translated into English during transcription, and a sample of transcripts was reviewed against the original recordings to verify translation accuracy. Data were analyzed using a hybrid inductive–deductive thematic approach informed by the TDF. Transcripts were read repeatedly to achieve familiarization, after which inductive coding was undertaken to identify barriers and facilitators to diabetes management. Emerging codes were grouped into categories and subsequently mapped to relevant TDF domains, while codes that did not align with the framework were retained to preserve context-specific insights.

A coding framework was developed and managed using NVivo version 15 (Lumivero, Denver, CO, USA). To enhance analytical rigor, a subset of transcripts was independently coded by the PI and research assistant, followed by consensus discussions to refine coding decisions and resolve interpretive differences. Themes were developed by examining patterns within and across participant groups. Comparisons between patient and healthcare provider perspectives were used to identify areas of convergence and divergence and to develop a comprehensive understanding of behavioral, social, and health-system influences on diabetes management.

### Quality assurance and trustworthiness

Several strategies were employed to enhance the rigor and trustworthiness of the study. Interview guides were piloted prior to data collection, and regular debriefing meetings were conducted to review interview quality and emerging findings. Credibility was strengthened through triangulation of patient and healthcare provider perspectives, transcript verification, and independent coding of selected transcripts. Dependability and confirmability were supported through maintenance of an audit trail documenting methodological and analytical decisions, while ongoing reflexive discussions helped examine potential sources of bias. Detailed descriptions of the study setting, participants, and research procedures were provided to facilitate assessment of the transferability of the findings. Reporting was guided by the Consolidated Criteria for Reporting Qualitative Research (COREQ).

## Results

### Demographic and socio-clinical characteristics of participants In-Depth Interview (IDI) participants profile

A total of 15 in-depth interviews were conducted with adult patients receiving diabetes care at Kasangati Health Centre IV. Participants ranged in age from 30 to 87 years, with the largest proportion aged 61–75 years (33.3%). Most participants were female (66.7%), and two-thirds lived with dependents (66.7%). More than half of the participants (53.3%) had lived with diabetes for over 10 years, while nearly half (46.6%) resided between 5 and 10 km from the health facility (Table 1).

**Table 1:** Socio-demographic characteristics of IDI participants (N=15)

| Variable | Frequency (n) | Percentage (%) |
| --- | --- | --- |
| **Age distribution (years)** |  |  |
| 30–45 | 4 | 26.7 |
| 46–60 | 4 | 26.7 |
| 61–75 | 5 | 33.3 |
| 76–87 | 2 | 13.3 |
| **Gender** |  |  |
| Female | 10 | 66.7 |
| Male | 5 | 33.3 |
| **Living status** |  |  |
| Living alone | 5 | 33.3 |
| Living with dependents | 10 | 66.7 |
| **Duration of illness (years)** |  |  |
| <5 | 3 | 20.0 |
| 5–10 | 4 | 26.7 |
| >10 | 8 | 53.3 |
| **Estimated distance to facility (km)** |  |  |
| <5 | 4 | 26.7 |
| 5–10 | 7 | 46.6 |
| >10 | 4 | 26.7 |

#### 1.2. Key Informant interview (KII) participants profile

Eight key informant interviews were conducted with healthcare and administrative personnel involved in diabetes service delivery at Kasangati Health Centre IV. Participants represented different professional cadres involved in clinical care, nursing services, laboratory diagnostics, pharmacy services, and facility management, providing diverse perspectives on diabetes care delivery and health system operations (Table 2).

**Table 2:**
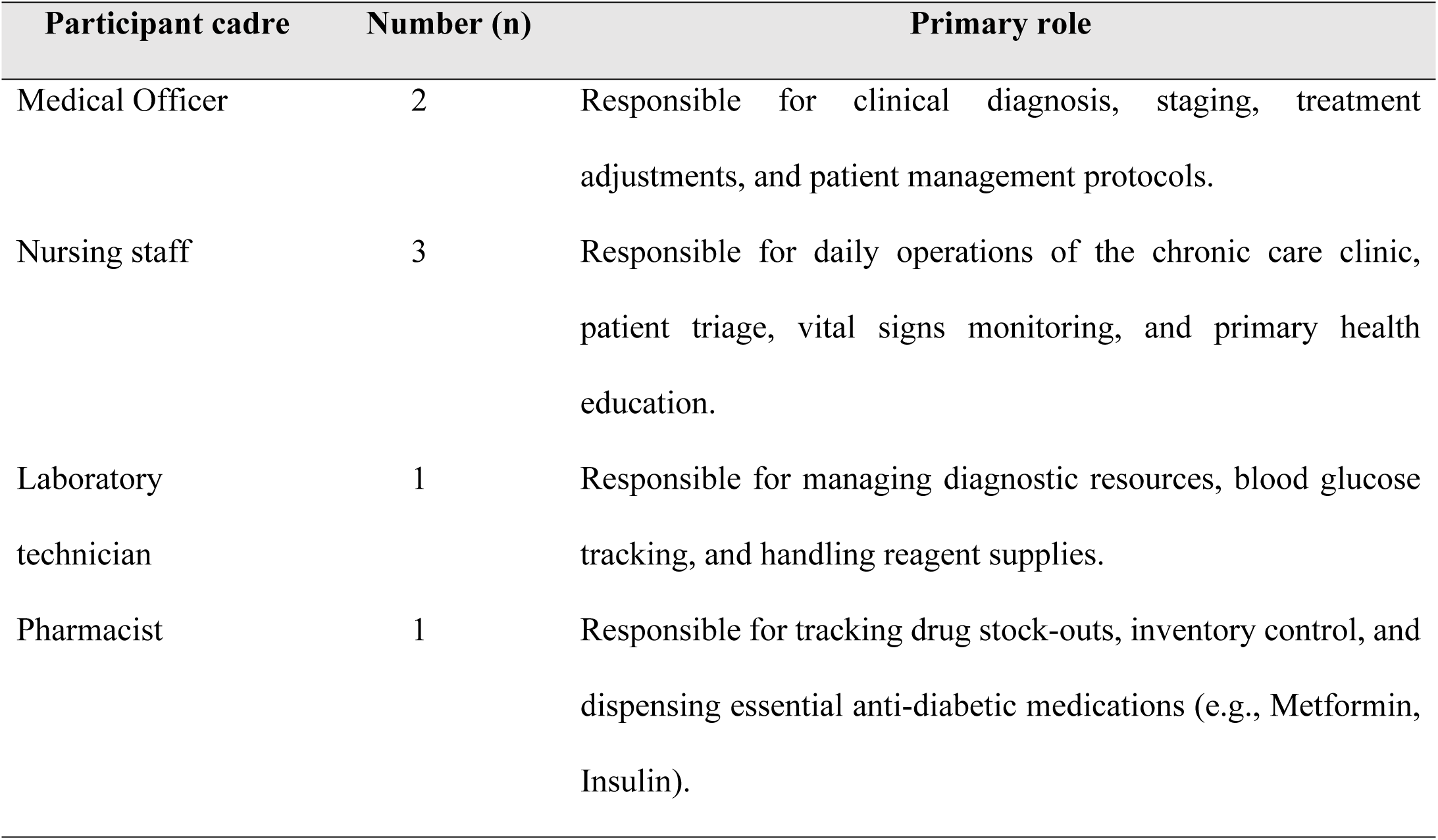
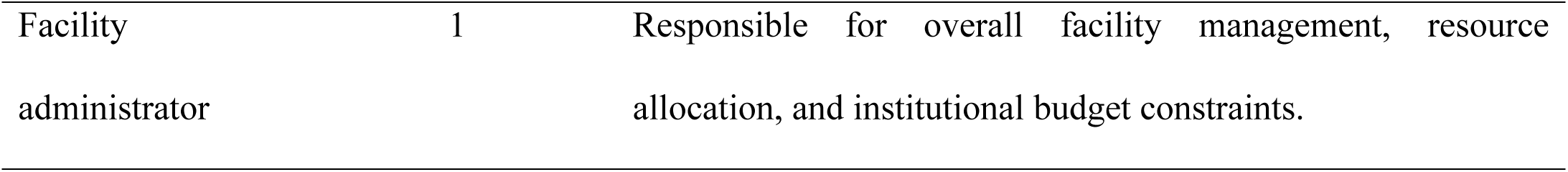
Professional profile of key informants (N=8)

#### Theme 1: Institutional and environmental influences on care access

Participants described several institutional and environmental factors that influenced access to and delivery of diabetes care at Kasangati Health Centre IV. Both patients and healthcare providers highlighted challenges related to physical infrastructure, communication systems, patient flow, and resource availability. Despite these barriers, participants also described adaptive strategies that facilitated access to care.

##### Physical access and communication challenges

Patients reported difficulties navigating the health facility, particularly older adults and those with mobility, visual, or hearing impairments. Long walking distances within the facility, inadequate seating, and challenges hearing queue announcements were commonly reported. Several participants also indicated that English-only signage limited their ability to identify service points and navigate the clinic independently.

> *“The walking distance from the gate to the clinic is difficult when your joints are painful, and there are often not enough places to sit.” (IDI-PT-04)*
>
> *“When you cannot read English, it is difficult to know where to go unless someone helps you.” (IDI-PT-07)*

##### Resource and infrastructure constraints

Healthcare providers acknowledged many of the challenges reported by patients and attributed them to broader resource and infrastructure limitations within the facility. Participants described high patient volumes, inadequate staffing, limited physical space, and the absence of communication aids such as public address systems as barriers to effective service delivery. These constraints were perceived to affect patient navigation, waiting times, and providers’ ability to offer individualized support.

> *“We do not have a public address system, so patients are called manually. Those with hearing difficulties can easily miss their turn.” (KII-HP-02)*
>
> *“The facility was designed for a smaller population, but demand for services has increased substantially. Our infrastructure and resources have not expanded at the same pace.” (KII-HP-08)*

##### Facilitators to care access

Despite these challenges, participants described several practices that helped facilitate access to care. Healthcare providers reported informally prioritizing frail or elderly patients when possible, while patients described relying on peer support from other clinic attendees to navigate clinic processes and receive assistance with queue management.

> *“The nurses sometimes allow very old or very sick patients to be seen earlier, which helps reduce the burden of waiting.” (IDI-PT-08)*
>
> *“I usually sit near someone who can hear better than me and ask them to let me know when my number is called.” (IDI-PT-01)*

#### Theme 2: Cognitive and informational factors influencing medication adherence

Participants described several challenges related to understanding, remembering, and implementing medication instructions. Both patients and healthcare providers identified limitations in health literacy, medication knowledge, and counseling processes that affected adherence to diabetes treatment. Despite these challenges, participants described strategies that supported medication management and continuity of care.

##### Knowledge gaps and medication management challenges

Patients reported difficulties understanding medication instructions, particularly when treatment regimens changed or when medications needed to be taken under specific conditions. Some participants described modifying medication use based on personal interpretations of symptoms, fear of adverse effects, or uncertainty regarding the purpose of prescribed medicines.

> *“Sometimes I skip the medicine when I have not eaten because I am afraid my sugar will go too low.” (IDI-PT-11)*
>
> *“When the doctors change the tablets, I get confused. If I feel dizzy, I sometimes think the medicine is too strong.” (IDI-PT-06)*

##### Counseling and information delivery constraints

Healthcare providers acknowledged challenges in delivering comprehensive medication counseling within a busy clinic environment. Participants reported that high patient volumes and limited staffing reduced opportunities for individualized education and follow-up. Providers also highlighted the lack of locally appropriate educational materials to reinforce medication instructions outside the clinical encounter.

> *“When many patients are waiting, counseling is often limited to brief instructions on how to take the medicine.” (KII-HP-04)*
>
> *“We do not have enough printed materials or visual aids that patients can take home and refer to later.” (KII-HP-03)*

##### Facilitators to medication adherence

Despite these barriers, participants identified several factors that supported medication adherence. Healthcare providers described conducting routine health education sessions before consultations, which helped reinforce key messages about diabetes management. Patients also reported developing practical strategies to remember medication schedules, including linking medication intake to daily routines and using household objects to distinguish between different medicines.

> *“The health talks before clinic help us understand how to manage our diabetes.” (IDI-PT-13)*
>
> *“I keep my medicines in different containers so that I can easily remember which one to take.” (IDI-PT-10)*

#### Theme 3: Social influences on diabetes management

Participants described how social relationships and community influences shaped diabetes management. While some patients experienced stigma, social isolation, and pressure to use traditional remedies, others reported receiving practical and emotional support from religious and community networks. Healthcare providers also highlighted challenges in addressing community-level influences on treatment adherence.

##### Stigma, social isolation, and traditional medicine influences

Several patients reported experiencing stigma and misconceptions about diabetes within their communities. Participants described situations where symptoms such as swelling, weakness, or chronic illness were attributed to witchcraft, curses, or other supernatural causes. These beliefs sometimes encouraged patients to seek traditional remedies or discontinue prescribed medications.

> *“Some neighbours told me my illness was caused by a curse and advised me to stop taking hospital medicine and seek traditional treatment.” (IDI-PT-12)*
>
> *“People told me that diabetes medicine would harm me and encouraged me to use local herbs instead. I stopped my medicine for some time and became seriously ill.” (IDI-PT-09)*

##### Limited community follow-up and support mechanisms

Healthcare providers acknowledged that community influences could affect treatment adherence but reported limited capacity to monitor patients outside the facility. Participants indicated that resource constraints restricted community outreach activities and reduced opportunities for ongoing engagement with patients and their families.

> *“Our services are mainly facility-based, and we have limited resources for community follow-up of patients with diabetes.” (KII-HP-01)*
>
> *“We know that patients receive different advice in their communities, but we have few opportunities to engage families or conduct follow-up visits.” (KII-HP-06)*

##### Community and religious support networks

Despite these challenges, participants described several forms of social support that facilitated diabetes management. Religious groups, friends, and community members were frequently mentioned as sources of emotional encouragement, transport assistance, and reminders to attend clinic appointments and adhere to treatment.

> *“Members of my church help me with transport when I cannot afford to come to the clinic.” (IDI-PT-03)*
>
> *“People from my mosque help me get to the clinic each month and encourage me to continue with treatment.” (IDI-PT-14)*

#### Theme 4: Emotional experiences of patients and healthcare providers

Participants described a range of emotional experiences associated with diabetes management and care delivery. Patients reported feelings of loneliness, worry, and discouragement related to living with a chronic illness, while healthcare providers described emotional strain associated with caring for large numbers of patients within a resource-constrained environment.

##### Emotional distress among patients

Several patients described feelings of loneliness and emotional distress associated with managing diabetes, particularly among those living alone or lacking regular family support. Participants expressed concerns about their health, future wellbeing, and ability to continue managing their condition over time. Some reported that emotional distress occasionally reduced their motivation to adhere to treatment recommendations.

> *“Sometimes I worry about what would happen if I became seriously ill while alone at home.” (IDI-PT-03)*
>
> *“Living with this disease alone can be discouraging, and there are times when you feel tired of following all the treatment instructions.” (IDI-PT-15)*

##### Emotional strain among healthcare providers

Healthcare providers reported emotional challenges associated with delivering diabetes care in a setting characterized by high patient volumes, staffing shortages, and periodic medication stockouts. Participants described frustration when they were unable to provide the level of care they believed patients required.

> *“It is difficult when patients come expecting medicines and we do not have them available.” (KII-HP-06)*
>
> *“Sometimes there are many patients waiting, and it becomes challenging to give each person the time and attention they need.” (KII-HP-03)*

##### Sources of emotional resilience

Despite these challenges, participants identified several factors that helped them cope with emotional stress. Healthcare providers described receiving support from colleagues through teamwork and informal discussions, while patients frequently cited faith, prayer, and personal determination as important sources of strength.

> *“We support each other as colleagues when the clinic becomes overwhelming.” (KII-HP-04)*
>
> *“Prayer and faith help me remain hopeful and continue taking my medicine.” (IDI-PT-07)*

#### Theme 5: Self-management strategies and continuity of care

Participants described several challenges and strategies related to maintaining treatment adherence and continuity of diabetes care. While healthcare providers highlighted limitations in patient follow-up systems, patients reported developing practical approaches to remember appointments, manage medications, and remain engaged in care.

##### Challenges in continuity of care

Healthcare providers reported that diabetes care at the facility relied largely on paper-based records and patient-initiated follow-up. Participants noted that the facility had limited capacity to track patients who missed appointments or to conduct routine follow-up outside the clinic setting.

> *“If a patient misses an appointment, we do not have a system to routinely contact them or follow them up at home.” (KII-HP-05)*
>
> *“Our records are mainly used for documentation and reporting, but they do not allow real-time tracking of patients who stop attending care.” (KII-HP-01)*

##### Patient strategies for self-management

Patients described developing practical methods to help them remember clinic appointments and medication schedules. These strategies were often adapted to individual circumstances and included linking medication-taking or clinic attendance to familiar daily routines, household objects, or recurring community events.

> *“I use events in the community to help me remember when it is time to return to the clinic.” (IDI-PT-11)*
>
> *“I keep a reminder near my water container so that I remember to take my medicine every morning.” (IDI-PT-02)*

##### Community support for continuity of care

Patients also described relying on trusted family members, neighbours, religious networks, and community members to support treatment adherence and clinic attendance. In some cases, community members assisted with transport, medication collection, or appointment reminders.

> *“A trusted boda-boda rider helps me collect my medicines when I am unable to travel to the clinic myself.” (IDI-PT-05)*

Across the themes, both patients and healthcare providers identified barriers and facilitators operating at individual, social, and health-system levels. While many perspectives converged, differences emerged in how the two groups understood the underlying causes of diabetes management challenges (Table 3).

**Table 3.**
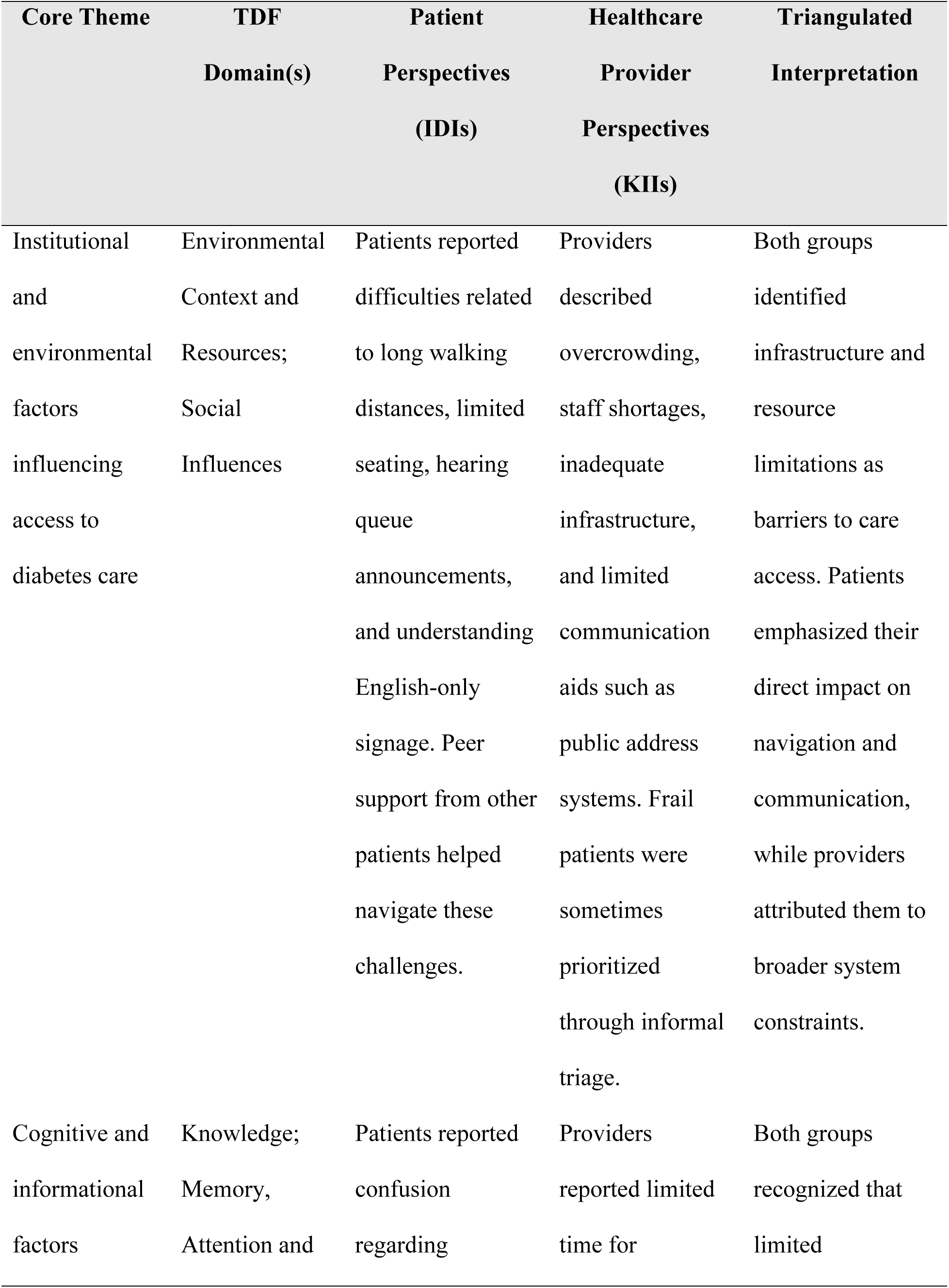
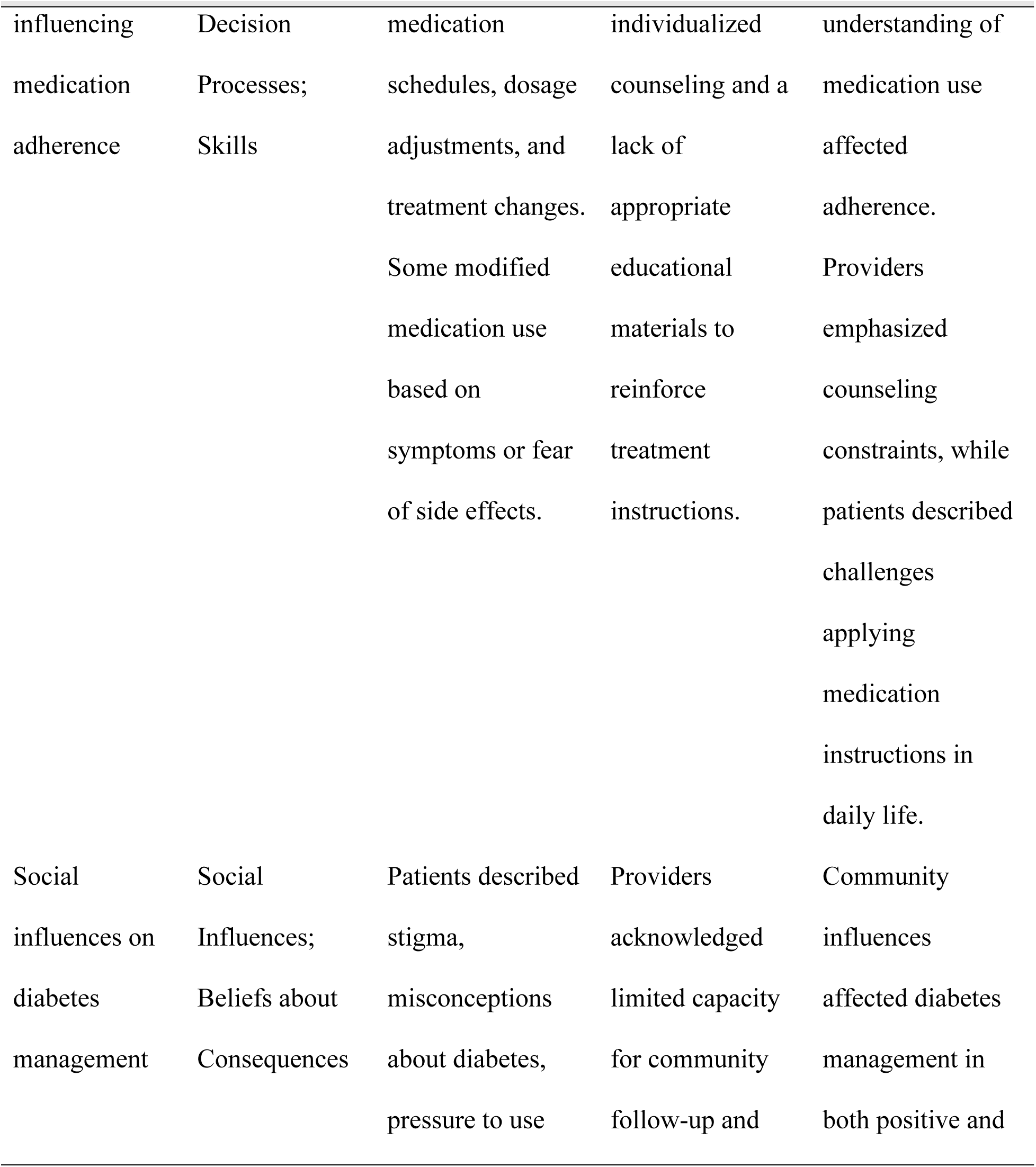
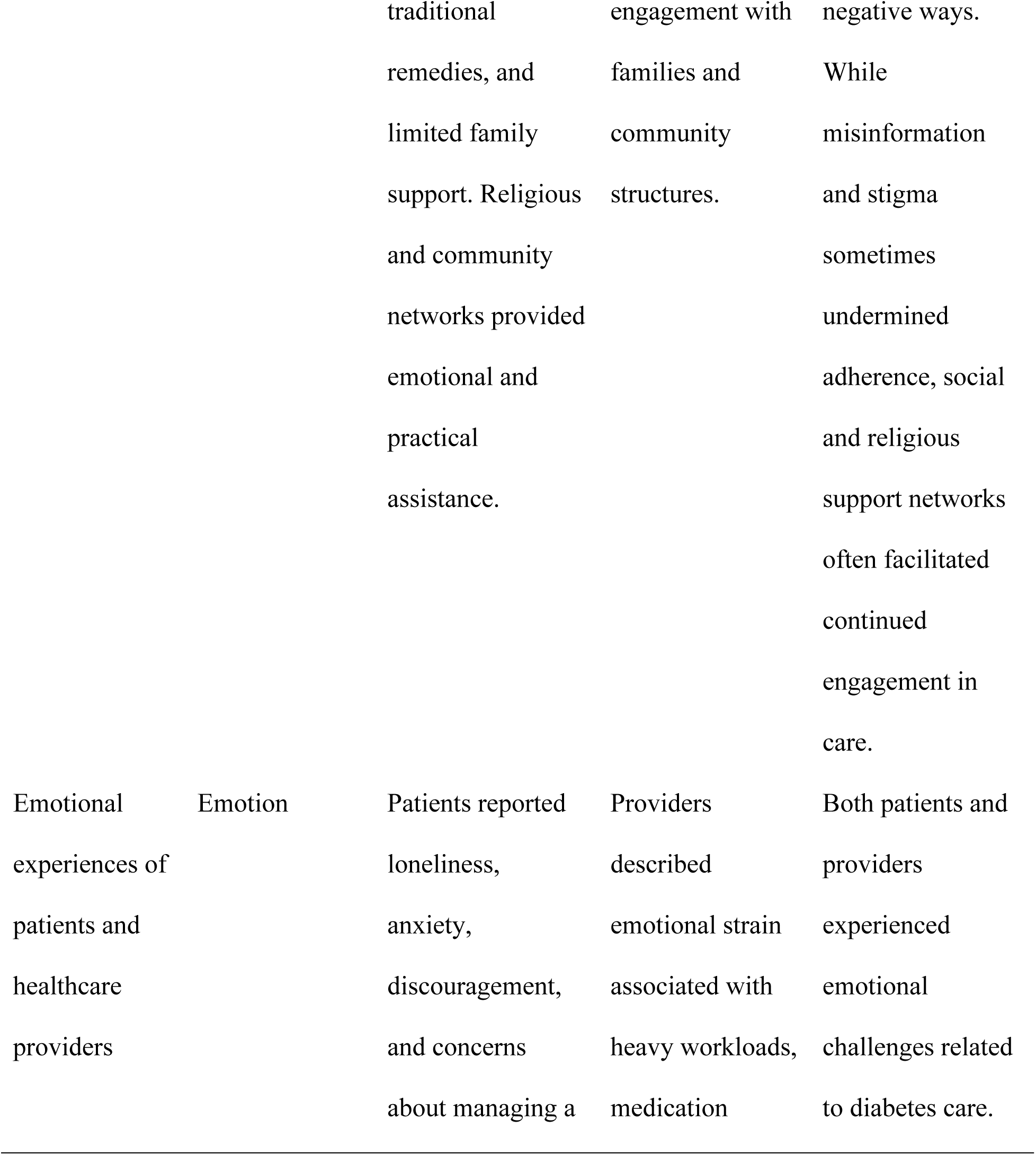
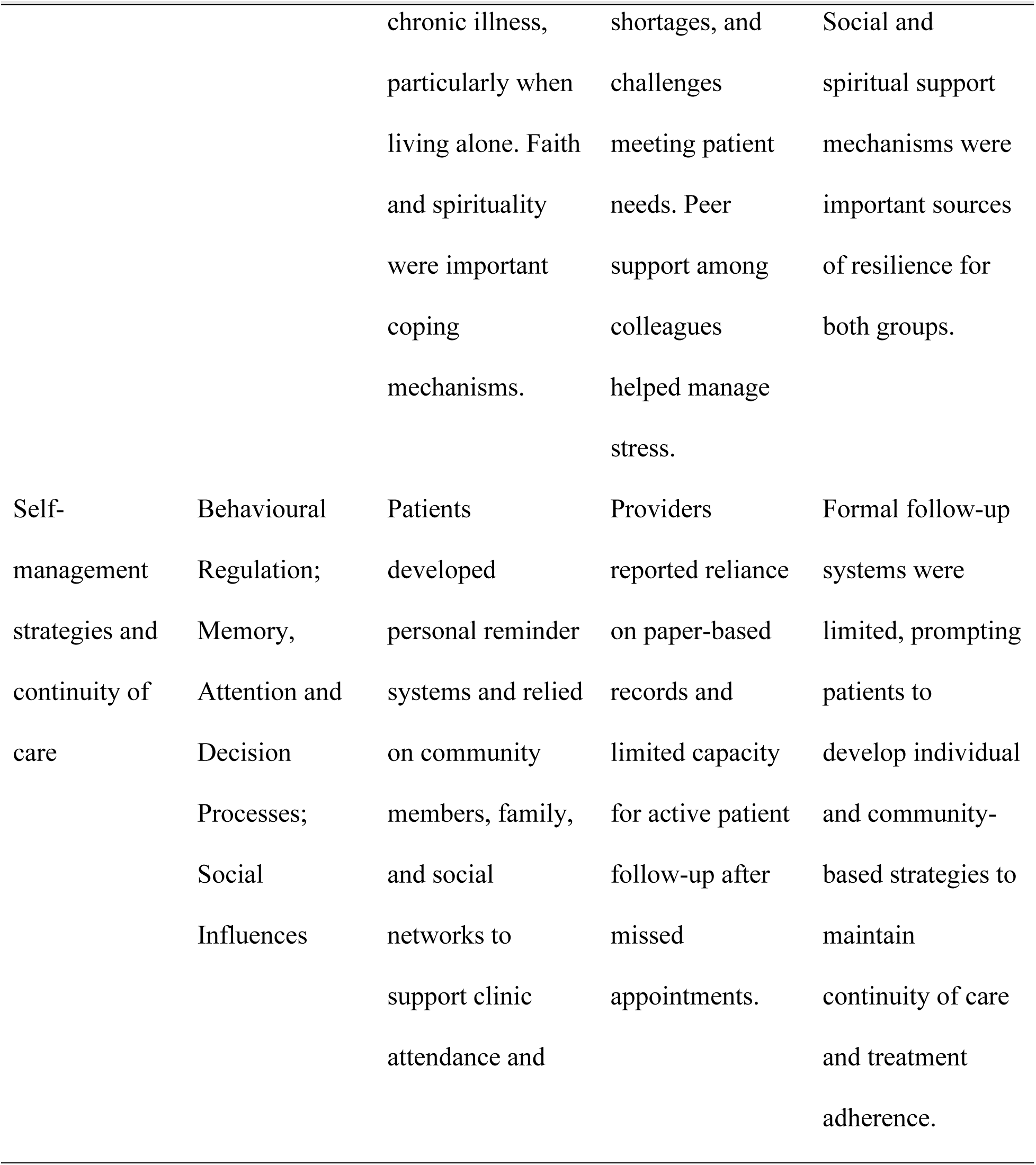
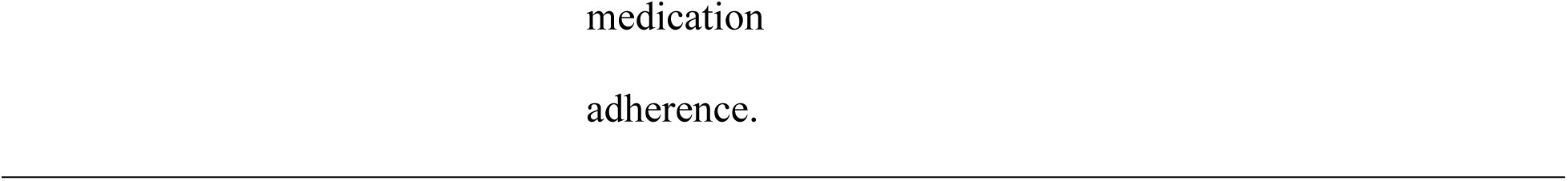
Triangulation matrix of patient and healthcare provider perspectives mapped to the theoretical domains framework.

## Discussion

In this study, barriers and facilitators to diabetes management were explored from the perspective of patients and healthcare providers at a peri-urban health facility in Uganda. It is evident from the results of this study that there are many factors, including system health factors, information-related factors, social factors, emotional factors, and behavior-related factors, that influence diabetes management. Some of the barriers mentioned by participants include limited resources, communication challenges, lack of health literacy, stigma, and lack of continuity of care programs, while some of the facilitators include social support, health education, provider support, and patient self-management strategies.

From the findings above, it is clear that diabetes care is very much affected by the ability of the health system to deliver accessible and sustained chronic care. Issues arising from the delivery environment could affect the patient’s ability to access the care consistently, hence indicating that diabetes care outcomes are determined not only by individuals but also by the care delivery environment. Such issues have been observed elsewhere in sub-Saharan Africa as health systems strive to deliver quality services amid the rising burden of non-communicable diseases in an acute health delivery environment [12, 13]. Just like the domain of environmental context and resources in the TDF [11], the above findings suggest a need for interventions aimed at enhancing the delivery environment through strengthening the health system. Efforts to address such constraints may be even more critical in peri-urban areas due to the rising demand for chronic care services. The findings indicate that medication adherence is influenced not only by access to medicines but also by patients’ understanding of treatment instructions and their ability to apply this information in everyday life. Participants described challenges interpreting medication guidance, remembering treatment schedules, and adapting recommendations to their daily routines. Similar barriers have been reported in studies from Uganda, Kenya, Ethiopia, and Tanzania, where limited health literacy and inadequate patient education have been associated with poor adherence and suboptimal diabetes outcomes [14, 15]. These findings correspond to the TDF domains of knowledge, skills, and memory, attention and decision processes, which emphasize the importance of understanding, retaining, and applying health information to support behavior change [11]. The findings suggest that diabetes self-management cannot be assumed solely on the basis of information provision. Rather, effective self-management requires communication approaches that account for varying literacy levels, language preferences, and cognitive demands associated with chronic disease management. This highlights the importance of patient education strategies that are accessible, culturally appropriate, and reinforced over time.

These results indicate the significance of social context in diabetes management, emphasizing the fact that self-management practices are often determined by the involvement of family members, friends, religious groups, and the larger community. Social support became one of the key factors in adherence since it offered emotional, practical, and reminder support regarding the treatment and visits to the clinic. On the other hand, the participants mentioned the influence of stigma associated with diabetes and other conflicting ideas regarding health. This dual role of social networks has been reported in other studies across sub-Saharan Africa, where families and communities often serve as important sources of support while also influencing health behaviours through prevailing beliefs and social expectations [15, 16]. Consistent with the TDF domain of Social Influences [11], these findings suggest that diabetes management is not solely an individual responsibility but is embedded within broader social contexts. Consequently, interventions that actively engage family members, peer-support groups, religious institutions, and community structures may strengthen adherence and improve long-term diabetes outcomes.

The experience of being affected emotionally by the challenge of dealing with diabetes was found to be a significant part of the process of management of this disease in this research. The majority of participants felt concerned, uncertain, frustrated, and exhausted emotionally in their efforts to deal with a life-long disease requiring constant monitoring. This kind of emotional challenges has been mentioned before in other research from sub-Saharan Africa, when emotional distress was considered a factor leading to decreased motivation, lack of compliance and disengagement in care [17, 18]. Interestingly enough, the emotional stress was not experienced only by patients. Healthcare professionals spoke about the difficulties they had to face due to the volume of patients they needed to deal with and the limited number of resources in the healthcare system, which made their work harder. The importance of the emotional state of both patients and providers is demonstrated by the fact that emotion plays an important role in the TDF [11]. The introduction of psychosocial support into regular diabetes management could prove beneficial for both patients and providers.

These results illustrate how patients actively tailor the diabetes treatment recommendations to fit the reality of their day-to-day lives. It seems that the maintenance of long-term care relied on more than just the accessibility of healthcare services, but was contingent on patients’ capacity to incorporate the therapy into their routine and utilize the social resources that were available to them. The same results have been observed throughout the sub-Saharan Africa where patients who suffer from chronic illnesses come up with practical coping strategies allowing them to stay committed to their care despite the presence of financial, transport-related, and health system-related difficulties [19, 20]. These results align with the domains of Behavioural Regulation, memory, attention and decision processes, and social influences of the TDF [11]. Thus, continuity of care is facilitated not only through individual self-regulation but also through social support systems. Improving the health outcomes of patients with diabetes may require interventions based on this pattern.

### Implications for policy and practice

The findings of this study suggest that improving diabetes management in peri-urban primary healthcare settings requires interventions that address patient-, community-, and health-system-level barriers simultaneously. The environmental and communication challenges identified by participants highlight the need for more accessible and patient-centered chronic care services, particularly for older adults and individuals with limited literacy. Health facilities may benefit from adopting context-appropriate communication strategies, including the use of local languages, visual educational materials, and simplified patient navigation systems.

The study also underscores the importance of strengthening diabetes self-management support. The mismatch between information provided by healthcare workers and patient understanding suggests a need for structured diabetes education that extends beyond routine clinical consultations. Educational interventions should be tailored to local literacy levels and reinforced through repeated engagement during follow-up visits.

At the community level, the findings demonstrate the influential role of social networks, religious institutions, and community beliefs in shaping diabetes management behaviors. Strengthening community-based support mechanisms, including engagement of Village Health Teams (VHTs), faith-based organizations, and peer-support networks, may help reinforce adherence, reduce stigma, and improve continuity of care.

Finally, the emotional burden experienced by both patients and healthcare providers highlights the need for more holistic chronic care approaches. Integrating psychosocial support into diabetes services and strengthening supportive workplace environments for healthcare providers may improve patient experiences and the quality of diabetes care. Collectively, these findings support the development of integrated, person-centered diabetes programs that address behavioral, social, and health-system determinants of chronic disease management.

### Strengths and Limitations

A key strength of this study was the inclusion of both patient and healthcare provider perspectives, which enabled a more comprehensive understanding of the factors influencing diabetes management within a peri-urban primary healthcare setting. The use of qualitative methods allowed for in-depth exploration of experiences that are often not captured through quantitative approaches alone. Additionally, the application of the Theoretical Domains Framework provided a structured approach for examining behavioral, social, and health-system influences on diabetes management, thereby enhancing the interpretability and potential applicability of the findings to intervention design.

Several limitations should be considered when interpreting the findings. First, the study was conducted at a single Health Centre IV in Wakiso District, which may limit the transferability of findings to other settings with different patient populations or health-system contexts. Second, participants were recruited purposively and therefore may not represent all individuals living with diabetes or all healthcare providers involved in diabetes care. Third, as with all interview-based studies, the findings may have been influenced by recall bias or social desirability bias. Nevertheless, efforts to enhance trustworthiness included the use of trained interviewers, triangulation of patient and provider perspectives, reflexive discussions during analysis, and adherence to established qualitative reporting standards.

## Conclusion

Diabetes management in this peri-urban Ugandan setting was influenced by a complex interplay of health-system, informational, social, emotional, and behavioral factors. The findings suggest that effective diabetes care requires approaches that extend beyond clinical treatment to address broader contextual influences on self-management and service utilization. While patients demonstrated considerable resilience in adapting to challenges associated with long-term disease management, persistent barriers within the health system and community environment may undermine sustained engagement in care. Strengthening diabetes outcomes will likely require multifaceted interventions that improve the accessibility and organization of services, enhance patient education, leverage community and social support structures, and integrate psychosocial support within routine diabetes care. The Theoretical Domains Framework proved useful in identifying modifiable determinants of diabetes management and may inform the development of contextually appropriate interventions for strengthening chronic disease care in similar resource-constrained primary healthcare settings.

## Declarations

### Availability of data and materials

The datasets generated and analyzed during the current study are not publicly available in full because the interview transcripts contain potentially identifiable information from study participants. De-identified excerpts supporting the findings are included within the manuscript and de-identified data may be made available from the corresponding author through Makerere University, school of Public Health Research Ethics Committee at.

### Competing interests

The authors declare that they have no competing interests.

### Consent for publication

Not applicable.

### Funding

The authors received no specific funding for this study.

### Author contributions

L.N.Y.K. and R.K. conceptualized the study, developed the methodology, conducted data collection, and drafted the initial manuscript. L.N.Y.K., R.K., and R.K. led the data analysis, coding, interpretation of findings, and preparation of tables and figures. D.M., R.K., O.N., M.W.W.D., R.K and A.D.K.N. provided supervision, contributed to study design refinement, critically reviewed the manuscript, and supported interpretation of the findings. All authors reviewed, revised, and approved the final manuscript.

## Acknowledgements

The authors thank the administration and staff of Kasangati Health Centre IV for their support during the study. We are also grateful to all patients and healthcare providers who generously shared their experiences and perspectives. Their participation made this study possible.

## Supporting Information

**S1 File. Interview guides**

Semi-structured interview guides used for in-depth interviews with patients and key informant interviews with healthcare providers.

## Notes

### Competing Interest Statement

The authors have declared no competing interest.

### Author Declarations

The study was conducted in accordance with the Declaration of Helsinki and approved by the Makerere University School of Public Health Research and Ethics Committee (MakSPH-REC_502). Administrative permission was obtained from Kasangati Health Centre IV (HCIV). Informed verbal consent was obtained from all participants before data collection. The use of verbal consent was approved by the Research Ethics Committee to minimize the collection of identifying information and enhance confidentiality. Participants received information about the study objectives, procedures, potential risks and benefits, voluntary participation, and confidentiality protections in their preferred language (Luganda for patients and English for healthcare providers). They were informed of their right to decline participation, refuse to answer any question, or withdraw from the study at any time without consequences. No personal identifiers were collected. Audio recordings and transcripts were assigned unique identification codes, and all identifying information was removed during transcription. Electronic data were encrypted and stored on password-protected devices accessible only to the research team.

## References

1. Federation ID. IDF Diabetes Atlas 11th Edition—Middle-East & North Africa (MENA) Fact Sheet. International Diabetes Federation. 2025.

2. World Health Organization. Global report on diabetes. Geneva: World Health Organization; 2016 [cited 2026 June 9]. Available from: https://www.who.int/publications/i/item/9789241565257

3. Atun R, Davies JI, Gale EA, Bärnighausen T, Beran D, Kengne AP, Levitt NS, Mangugu FW, Nyirenda MJ, Ogle GD, Ramaiya K. Diabetes in sub-Saharan Africa: from clinical care to health policy. The lancet Diabetes & endocrinology. 2017 Aug 1;5(8):622–67.

4. Pastakia SD, Pekny CR, Manyara SM, Fischer L. Diabetes in sub-Saharan Africa–from policy to practice to progress: targeting the existing gaps for future care for diabetes. Diabetes, metabolic syndrome and obesity: targets and therapy. 2017 Jun 22:247–63.

5. World Health Organization. Uganda STEPS survey 2014: fact sheet. Geneva: World Health Organization; 2014 [cited 2026 June 9]. Available from: https://cdn.who.int/media/docs/default-source/ncds/ncd-surveillance/steps/uganda-steps-2014-factsheet.pdf

6. Kusolo R, Mutungi GN, Mbuliro M, Kajjura R, Wesonga R, Bahendeka SK, Guwatudde D. Changes in the prevalence of the common risk factors for non-communicable diseases in Uganda between 2014 and 2023: Informed by nationally representative cross-sectional surveys. PLOS Global Public Health. 2025 Apr 8;5(4):e0003755.

7. Bahendeka S, Wesonga R, Mutungi G, Muwonge J, Neema S, Guwatudde D. Prevalence and correlates of diabetes mellitus in Uganda: a population-based national survey. Tropical Medicine & International Health. 2016 Mar;21(3):405–16.

8. Birabwa C, Bwambale MF, Waiswa P, Mayega RW. Quality and barriers of outpatient diabetes care in rural health facilities in Uganda–a mixed methods study. BMC Health Services Research. 2019 Oct 16;19(1):706.

9. Chang H, Hawley NL, Kalyesubula R, Siddharthan T, Checkley W, Knauf F, Rabin TL. Challenges to hypertension and diabetes management in rural Uganda: a qualitative study with patients, village health team members, and health care professionals. International journal for equity in health. 2019 Feb 28;18(1):38.

10. Makonje R, Omolo RO, Mudenda S, Mugenyi N. Prevalence and Factors Associated with Type 2 Diabetes Self-Management Among Patients Attending Mbale Regional Referral Hospital: A Cross-Sectional Study in Eastern Uganda. Diabetes, Metabolic Syndrome and Obesity. 2026 Dec 31:598398.

11. Cane J, O’Connor D, Michie S. Validation of the theoretical domains framework for use in behaviour change and implementation research. Implementation science. 2012 Apr 24;7(1):37.

12. Mercer T, Chang AC, Fischer L, Gardner A, Kerubo I, Tran DN, Laktabai J, Pastakia S. Mitigating the burden of diabetes in Sub-Saharan Africa through an integrated diagonal health systems approach. Diabetes, Metabolic Syndrome and Obesity. 2019 Oct 31:2261–72.

13. Kibirige D, Lumu W, Jones AG, Smeeth L, Hattersley AT, Nyirenda MJ. Understanding the manifestation of diabetes in sub Saharan Africa to inform therapeutic approaches and preventive strategies: a narrative review. Clinical diabetes and endocrinology. 2019 Feb 14;5(1):2.

14. Muwanguzi M, Obua C, Maling S, Wong W, Owokuhaisa J, Wakida EK. Barriers and facilitators to cognitive impairment screening among older adults with diabetes mellitus and hypertension by primary healthcare providers in rural Uganda. Frontiers in Health Services. 2023 May 30;3:1172943.

15. Salimu SN, Taylor M, A. Spencer S, Desmond N, Nyirenda D, Morton B. Self-management of chronic conditions including multimorbidity in sub-Saharan Africa: A systematic and meta-synthesis review with focus on diabetes, hypertension, chronic kidney disease, and HIV. PLOS Global Public Health. 2025 Oct 9;5(10):e0003836.

16. Onyango JT, Namatovu JF, Besigye IK, Kaddumukasa M, Mbalinda SN. The relationship between perceived social support from family and diabetes self-management among patients in Uganda. The Pan African Medical Journal. 2022 Apr 7;41:279.

17. Mendenhall E, Musau A, Bosire E, Mutiso V, Ndetei D, Rock M. What drives distress? Rethinking the roles of emotion and diagnosis among people with diabetes in Nairobi, Kenya. Anthropology & Medicine. 2020 Jul 2;27(3):252–67.

18. Kalra S, Jena BN, Yeravdekar R. Emotional and psychological needs of people with diabetes. Indian journal of endocrinology and metabolism. 2018 Sep 1;22(5):696–704.

19. Masupe T, Onagbiye S, Puoane T, Pilvikki A, Alvesson HM, Delobelle P. Diabetes self-management: a qualitative study on challenges and solutions from the perspective of South African patients and health care providers. Global Health Action. 2022 Dec 31;15(1):2090098.

20. Desse TA, Namara KM, Manias E. Patient-perceived challenges to type 2 diabetes self-management in sub-Saharan Africa: a qualitative exploratory study. The Science of Diabetes Self-Management and Care. 2024 Dec;50(6):456–68.

